# A randomised controlled trial of preconception lifestyle intervention on maternal and offspring health in people with increased risk of gestational diabetes: study protocol for the BEFORE THE BEGINNING trial

**DOI:** 10.1101/2023.07.18.23292734

**Authors:** MAJ Sujan, HS Skarstad, G Rosvold, SL Fougner, SA Nyrnes, AC Iversen, T Follestad, KÅ Salvesen, T Moholdt

**Affiliations:** Department of Circulation and Medical Imaging, Norwegian University of Science and Technology, Trondheim, Norway; Department of Women’s Health, St. Olavs Hospital, Trondheim University Hospital, Trondheim, Norway; Department of Clinical and Molecular Medicine, Norwegian University of Science and Technology, Trondheim, Norway; Department of Endocrinology, St. Olavs Hospital, Trondheim University Hospital, Trondheim, Norway; Children’s clinic, St. Olavs Hospital, Trondheim University Hospital, Trondheim, Norway; Centre of Molecular Inflammation Research, Norwegian University of Science and Technology, Trondheim, Norway; Clinical Research Unit Central Norway, St. Olavs Hospital, Trondheim Norway

**Keywords:** insulin resistance, time-restricted eating, aerobic exercise, glycaemic control, diet

## Abstract

**Introduction:** Gestational diabetes mellitus (GDM) is associated with increased risk for type 2 diabetes in the mother and cardiometabolic diseases in the child. The preconception period is an optimal window to adapt the lifestyle for improved outcomes for both mother and child. Our aim is to determine the effect of a lifestyle intervention, initiated before and continued throughout pregnancy, on maternal glucose tolerance and other maternal and infant cardiometabolic outcomes.

**Methods and analysis:** This ongoing randomised controlled trial has included 167 females aged 18-39 years old at increased risk for GDM who are contemplating pregnancy. The participants were randomly allocated 1:1 to an intervention or control group. The intervention consists of exercise (volume is set by a heart rate-based app and corresponds to ≥ 1 hour of weekly exercise at ≥ 80% of individual heart rate maximum), and time-restricted eating (≤ 10 hours/day window of energy intake). The primary outcome measure is glucose tolerance in gestational week 28. Maternal and offspring outcomes are measured before and during pregnancy, at delivery, and at 6-8 weeks postpartum. Primary and secondary continuous outcome measures will be compared between groups based on the “intention to treat” principle using linear mixed models.

**Ethics and dissemination:** The Regional Committees for Medical and Health Research Ethics in Norway has approved the study (REK 143756). The anonymised results will be submitted for publication and posted in a publicly accessible database of clinical study results.

**Strengths and limitations of this study:**

- The intervention starts before and continues throughout pregnancy to make it easier for the participants to adopt an active lifestyle before pregnancy.
- This study includes individuals at high risk of GDM from multiple ethnic backgrounds, which improves the generalisability of the findings.
- The effects of the intervention on the cardiac function and body composition of the offspring will be comprehensively evaluated.
- Due to the difficulty of blinding investigators and participants to behavioural interventions, investigators will not be blinded for outcome assessments.
- Due to the long duration of the intervention, adherence to lifestyle modifications may be difficult for some participants despite regular monitoring and motivational support.

## Introduction

The global prevalence of gestational diabetes mellitus (GDM), i.e., high plasma glucose first identified during pregnancy, continues to increase. Both environmental and genetic factors contribute to the development of GDM, and up to 14 % of live births are negatively impacted by this condition.(1) GDM typically occurs because of pancreatic β-cell dysfunction with pre-existing insulin resistance and increases the risk for type 2 diabetes and cardiovascular disease in the mother.(2, 3) Maternal obesity and hyperglycaemia affect the offspring through the egg cell quality, intrauterine environment, and foetal organ development. These metabolic conditions eventually increase the risk for cardiac dysfunction at birth, and early onset diabetes, obesity, and cardiovascular diseases later in life.(4–9) Higher maternal blood glucose concentration, even below the diagnostic criteria for GDM, is associated with increased birth weight, elevated levels of cord-blood C-peptide, childhood obesity, and elevated blood pressure, independent of maternal body mass index (BMI).(10–12) Besides the inheritable risk factors, epigenetic modifications in utero, low-grade inflammation, and modifications of the gut microbiome can also negatively affect the cardiometabolic health of the offspring.(4)

Lifestyle interventions, including dietary changes, increased physical activity, and self-monitoring of blood glucose, are the first-line choice for GDM management.(13) However, many pregnant individuals fail to adhere to the recommendations for diet and exercise training(14, 15) and there is inconclusive evidence for clinically meaningful effects of diet-exercise interventions on pregnancy outcomes for the mother or child.(5, 16–18) Several recent randomised controlled trials (RCTs) and reviews conclude that pre-pregnancy lifestyle interventions are urgently needed to improve maternal health and increase the likelihood of adherence to a healthy lifestyle during pregnancy.(19–23) Alternative diet-exercise strategies, such as time-restricted eating (TRE) and high-intensity interval training (HIIT), have shown promising results on improving metabolic health among reproductive-aged females.(24–26) TRE is a safe and feasible intervention in individuals with overweight, obesity, prediabetes, and type 2 diabetes.(27) It has been shown to improve glucose tolerance, and insulin sensitivity, and reduce appetite, hunger, HbA1c, and total body and fat mass in this population.(28–36) While data on the effects of TRE in pregnancy are scarce, observational data suggest that longer maternal night-fasting intervals are associated with decreased fasting glucose.(37) The safety of HIIT is not yet established during pregnancy, but recent publications indicate that HIIT is safe, with higher enjoyment and improved adherence than continuous moderate-intensity training,(38) and may provide cardiometabolic benefits for both mothers and their offspring.(39–41)

Pre-pregnancy patterns of physical activity and exercise are important determinants of exercise during pregnancy,(42) and pre-pregnancy inception of healthy dietary habits is associated with a lower risk of GDM.(43–45) So far, there is limited evidence on the effectiveness of implementing both dietary and exercise-based lifestyle interventions before pregnancy. It is highly relevant to find feasible and effective pre-pregnancy lifestyle interventions which can reduce maternal hyperglycaemia and its related negative consequences for mother and child.

The primary hypothesis for the BEFORE THE BEGINNING (BTB) trial is that the participants allocated to the intervention group (time-restricted eating and exercise) will have improved maternal glucose tolerance in gestational week 28, compared with participants in the control group. We will also determine the effect of the intervention on secondary cardiometabolic outcomes in both the mothers and their newborns.

## Aims

### The primary aim of BEFORE THE BEGINNING

- To determine the effect of a lifestyle intervention, commenced preconception and continuing throughout pregnancy, on maternal glucose tolerance in pregnancy.

### Secondary aims of BEFORE THE BEGINNING

- To evaluate the effect of the intervention on insulin sensitivity, blood glucose, circulating lipids, body composition, cardiorespiratory fitness, systemic inflammation, and blood pressure in the mothers.
- To evaluate the effect of the intervention on cardiac function, body composition, and systemic inflammation in the newborns.
- To evaluate the adherence to the interventions, and their effects on sleep quality, appetite and hunger, physical activity, and dietary intake.

## Methods

### Design and study setting

This is an ongoing single-centre RCT with two parallel groups: an intervention group and a control group, undertaken at the Norwegian University of Science and Technology (NTNU) in Trondheim, Norway, in collaboration with the St. Olav’s Hospital, Trondheim, Norway. SPIRIT reporting guidelines were used in reporting this study protocol.(46)

### Recruitment and participants

The trial was announced through social media, hospital and university webpages, local stores, and public places. Additionally, potential participants were identified through the National Population Register, and we regularly sent out electronic invitations to females aged 20-35 years in Trondheim and the surrounding area to participate in the trial. The invitation prompted them to visit the study website, which contains a short description of the trial and allows potential participants to self-screen for eligibility before further screening by telephone. The first participant was included on 25^th^ September 2020 and the last participant was included on 28^th^ April 2023.

Box 1 shows the inclusion and exclusion criteria for participation in the study.

#### Box 1

Inclusion and exclusion criteria

##### Inclusion criteria

- Female
- Age: 18-39 years old
- Contemplating pregnancy within the next six months
- Understands oral and written Norwegian or English
- At least one of the following criteria must apply:
  - Body mass index ≥ 25 < 40 kg/m,
  - Gestational diabetes in a previous pregnancy,
  - Close relative with diabetes (either parents, siblings, or children with diabetes),

- Fasting plasma glucose > 5.3 mmol/L,
- Previous newborn > 4.5 kg, or
- Non-European ethnicity (with one or both parents originating from an area outside Europe).

##### Exclusion criteria

- On-going pregnancy
- Trying to conceive ≥ 6 cycles at study entry
- Known diabetes (type 1 or 2)
- Shift work that includes night shifts > 2 days per week
- Previous hyperemesis
- Known cardiovascular diseases
- High-intensity exercise > 2 times per week in the last 3 months
- Habitual eating window ≤ 12 hours
- Bariatric surgery
- Any other reason which according to the researchers makes the potential participant ineligible

### Randomisation and allocation

After screening and assessments at baseline, the participants were randomly allocated (1:1) to the intervention or a standard care control group, after stratifying for GDM in a previous pregnancy (yes/no). At the first visit, the study procedures, equipment, and applications were set up and explained to the participants. We used a computer random number generator (WebCRF3) developed and administered at The Clinical Research Unit (Klinforsk), NTNU/St. Olav’s Hospital, Trondheim, Norway to randomly allocate participants using various block sizes.

### Intervention

The intervention starts before pregnancy and continues throughout pregnancy and consists of a combination of TRE and exercise. Participants are counselled to change their daily time-window of energy intake to ≤ 10 hours, ending no later than 19:00 hours, for minimum 5 days per week throughout the intervention. The remaining 2 days are “days off” when they can consume food *ad libitum* if they wish. Apart from current recommendations about preconception/pregnancy nutrition, we give no advice regarding food choices, nor do we encourage a reduced total energy intake.

We use the Personal Activity Intelligence (PAI) score, a science-backed activity metric based on heart rate (HR)(47, 48) to prescribe exercise. Since PAI is HR-based, high-intensity exercise gives substantially higher PAI scores than low-to-moderate-intensity exercise. The goal for the participants in the intervention group is to earn and maintain ≥ 100 PAI per week, which can be reached by minimum 1 hour of weekly exercise at ≥ 80% of HR maximum. One week after the baseline visit, we invite the participants for a supervised introductory exercise session and provide a brochure with exercise options (e.g., treadmill walking/running, cycling). We invite the participants for a second session 2 weeks after the introductory session. The participants can choose their mode of exercise. Once pregnant, we advise the participants to either do short work-bouts at high intensity with low-to-moderate intensity periods in-between, or longer work periods up to 85% of heart rate maximum. We contact the participants not reaching 100 PAI to offer additional supervised exercise sessions, and they can also ask for extra support and supervised exercise sessions if they want to.

Participants in the control group receive standard care and are asked to continue with their habitual physical activity and dietary habits. We contact these participants once every 8 weeks to support adherence to registrations and monitoring.

### Experimental procedures and outcome measures

The study period spans from baseline assessments in the pre-pregnancy period to 6-8 weeks after delivery (Figure 1). Participants who do not become pregnant within 6 months after inclusion in the trial (changed from 12 months from December 2022, see below under modifications to the protocol after trial commencement) are excluded. For participants who experience spontaneous abortions, we add the number of weeks that the participant was pregnant plus 4 weeks to their time in the trial before exclusion pre-pregnancy.

**Figure 1.** Consort flow diagram of the BEFORE THE BEGINNING trial (Ongoing study: status 25.09.2020 – 17.07.2023) * If the participants are not pregnant within 12 months of inclusion, they are excluded from the study. From December 2022, the time-window for exclusion if not pregnant was reduced from 12 to 6 months.

Assessments of the participants are performed twice during preconception (at baseline before randomisation, and after 8 weeks), and twice during pregnancy (in gestational weeks 12 and 28). Outcomes in the newborns are assessed within 72 hours after delivery and at age 6-8 weeks (Figure 2). All participants receive a brochure from the Norwegian Health Directorate with the current recommendations for physical activity, diet, and folic acid, and iodine supplements. The participants are invited to ultrasound examinations in gestational weeks 12, 19, and 32.

**Figure 2.** Overview of time-points for assessments in the trial.

#### Primary outcome measure

The primary outcome measure is plasma glucose concentration obtained 2 hours after a 75 g oral glucose tolerance test (OGTT) in gestational week 28. After an overnight fast (≥ 10 hours) and no exercise for ≥ 24 hours, the participants consume 75 g of glucose (Glucosepro, Finnamedical, Finland) diluted in 250 mL water within 5 minutes. Using an indwelling catheter, we collect venous blood before the OGTT, with subsequent collections at 30, 60, 90, and 120 minutes after ingestion of glucose.

#### Secondary outcome measures

Secondary maternal and neonatal outcome measures (Figure 2) are described below.

#### Blood sampling and biochemistry

From all visits, fasting blood lipids, plasma glucose, and HbA1c are measured immediately after sampling, at St. Olav’s Hospital, following local standardised procedures. Additional fasting plasma, serum, full blood, and urine are stored in a biobank at -80°C for later analyses. GDM is recorded at Visit 3 and 4, according to the WHO 2013 criteria (fasting plasma glucose 5.1-6.9 mmol/L and/or 2-hour plasma glucose 8.5-11.0 mmol/L after 75 g OGTT).(49) At the event of a GDM diagnosis, the participant and their general practitioner are informed for further evaluation and management. Insulin sensitivity will be calculated using homeostasis model assessment of insulin resistance (HOMA-IR)(50) and pancreatic beta cell function using HOMA-β.(50) At visit 3 and 4, the area under the curve (AUC) and incremental AUC (iAUC) from glucose and insulin concentrations will be calculated from venous blood sampling every 30 minutes during the 2-hour OGTT. Insulin Sensitivity Index, ISI_0,120_,(51) insulinogenic index during the first 30 minutes of the 2-hour OGTT,(52) and beta cell function (AUC_ins_/AUC_glu_) will be estimated.(53)

#### Continuous glucose monitoring

The participants wear a continuous glucose monitor (CGM, FreeStyle Libre 1, Abbott Diabetes Care, Norway) for 14 days at baseline (7 days pre-intervention followed by the first 7 days of intervention/control), and for 14 days starting at 8 weeks from baseline. From these measurements, we will determine 24-hour glycaemic control, 3-hour postprandial glucose levels (AUC) for the first meal of the day, and nocturnal glycaemic control. The screens of the CGM readers are taped over to avoid lifestyle changes based on the participants’ glucose levels. We also plan to explore other CGM data that can predict glycaemic control during pregnancy, using machine learning.

#### Height, weight, body composition, BMI, and waist circumference

Height is measured with the participants standing without shoes using a standard stadiometer. Weight and body composition are estimated in the morning after overnight fasting using bioelectrical impedance analysis (Inbody 720, Biospace CO, Korea), with participants wearing light clothes and standing barefoot. BMI is calculated as weight in kilograms divided by the square value of height in metres (kg/m^2^). To account for the increase in fat-free mass hydration as pregnancy progresses, we will use a regression equation that estimates fat-free mass density as a function of gestational age.(54) Waist circumference is measured using a measuring tape at the level of the belly button with the participant standing.

#### Cardiorespiratory fitness

We measure peak oxygen uptake (VO_2peak_) using indirect calorimetry (Metalyzer II, Cortex, Germany), using an individualised treadmill protocol in which the participants walk or run until volitional exhaustion. The test starts after a 10-minute warm-up. The speed or inclination is increased every 1 – 2 minutes, by 0.5 – 1.0 km/hour or 1% – 2%. VO_2peak_ is determined as the average of the three highest consecutive 10 seconds measured and will be reported as both absolute (L/min) and relative (mL/min/kg) values. We record HR throughout the exercise tests and use the peak HR recorded during the test as an estimate of the HR maximum.(55)

#### Blood pressure and resting heart rate

We use an automatic blood pressure device (Welch Allyn, Germany) to measure blood pressure (diastolic and systolic, in mmHg) and resting HR (beats per minute, bpm) on the participants’ left arm after they have rested in a seated position for 15 minutes. We will report the average of three measurements taken at 1-minute intervals.

#### Physical activity, diet, and sleep

We use activity monitors to estimate physical activity levels, energy expenditure, and sleep duration. All participants wear Sensewear Armbands (BodyMedia, Pennsylvania, USA) for 14 days at baseline (7 days pre-intervention followed by the first 7 days of intervention/control), and the participants in the intervention group wear Amazfit GTS (Huami, China) smartwatches throughout the intervention. The smartwatch is connected to the Zepp app and shares PAI data with the research team via the Memento app. Participants register their diet in an online food diary (Fatsecret app) and record the time of first and last energy intake in the project handbook for 4 days (3 weekdays and 1 weekend day) every 8 weeks. They also complete questionnaires about physical activity, sleep quality, and psychological well-being every 8 weeks throughout the study period. We use the following questionnaires: 1) International Physical Activity Questionnaire,(56) 2) Pittsburgh Sleep Quality Index,(57) and 3) Psychological General Well-Being Index.(58) At baseline, the participants fill in the Horne-Östberg Morningsness-Eveningness Questionnaire.(59) We record medication and supplements, early miscarriages, abortions, and time to pregnancy and live birth. Additionally, expectant fathers are asked to complete questionnaires at baseline and every 8 weeks throughout the trial, including questions regarding their body weight, height, physical activity, and diet. These data will be used as co-variates in later analyses.

#### Neonatal and other outcomes

We obtain standard clinical neonatal outcomes from hospital birth records. Midwives at St. Olav’s Hospital collect umbilical cord blood immediately after birth, prior to the delivery of the placenta. Placental tissues are collected from 1) around the base of the umbilical cord on the foetal side, 2) the periphery on the maternal side (full-thickness tissue), 3) the centre of the maternal side, also for storage in RNAlater solution (Invitrogen, Thermofisher scientific, Lithuania) and 4% formaldehyde solution, and 4) the periphery on the maternal side. The samples are put in 1.8 mL cryotubes and snap-frozen immediately in liquid nitrogen, before storage at -80°C for later analyses. The samples in RNAlater solution are stored at 4°C overnight, followed by storage at -80°C for later analyses. The samples in 4% formaldehyde solution are stored under a fume hood at room temperature for 48 hours before histology slide preparation in collaboration with the CMIC Histology Lab at NTNU.

Within 72 hours of birth, and at age 6-8 weeks, body composition of the newborn is estimated using bioimpedance (BioScan touch i8-nano, Maltron, UK). Additionally, an experienced paediatric cardiologist examines cardiac morphology, structure, and function in the newborn, using a Vivid E95 scanner (GE Vingmed Ultrasound, Horten, Norway) and a GE 6s, and M5s phased-array transducers (GE Healthcare, Milwaukee, WI). A full clinical echocardiography including conventional echocardiographic parameters as well as study images with a focus on measurement of systolic and diastolic myocardial function is performed. The scanner is equipped with research software enabling high frame rate echocardiography to study cardiac flow and tissue properties as described previously.(60–62) A corresponding group of neonates (N = 30), from mothers with no known increased risk of GDM and BMI in the normal range (18.6-24.9 kg/m^2^) will be used for comparison.

#### Adherence

We record adherence to TRE as the average daily time-window for energy intake for 4 days every 8 weeks. Additionally, we categorise participants as adherent if they report a ≤ 10-hour time-window for energy intake on ≥ 2 of these 4 days. Adherence to exercise is recorded as the number of PAI points the participants get per rolling 7 days. To ensure compliance and maintain adherence, we send text messages to all participants as reminders to complete questionnaires and dietary reporting. We also announce friendly competitions such as “Who can keep 100 weekly PAI points or more for a whole month?” in a Facebook group for the participants. The data are only accessible to the researchers and a gift card is awarded to the winners.

### Modifications to the protocol after trial commencement

Since June 2021, we invite the participants to participate in a follow-up study after delivery in which we collect infant faecal samples (immediately after birth, at 6 weeks, and 6 months), maternal faecal samples (at 6 weeks and 6 months), and breast milk (at 6 weeks and 6 months). These samples are stored at -80°C for later analyses. Additionally, we started to offer supervised exercise training sessions to the participants in the intervention group. From November 2022, we started sending invitations using eFORSK (electronic form-based data collection, developed by Central Norway Regional Health Authority) and added ‘Bariatric surgery’ to the exclusion criteria. ‘Any other reason which according to the researchers makes the potential participant ineligible’ to undergo either or both interventions (e.g., traumatic foot injury, anorexia/bulimia, etc.) was also added to the exclusion criteria in November 2022. From December 2022, we removed ‘Planned assisted fertilisation with female factor reason’ from the exclusion criteria. In addition, we changed the maximum time before pregnancy from 12 months to 6 months to allow for the trial to be terminated in time for us to analyse the data within the project period. In March 2023, the required number of total participants was reduced from 260 to 200 based on the revised calculation as described in the sample size calculation, with additional specification of stopping before 200 if we had sufficient pregnant participants for the primary outcome measure. In June 2023, we changed from Amazfit GTS to Polar Ignite 2 (Polar, Finland) smartwatch and from Zepp and Memento to Polar Flow and Mia app.

### Sample size calculation

The primary outcome of this study is glucose tolerance (after a 2-hour OGTT) in gestational week 28. The HAPO study results(63) indicate strong, continuous associations of maternal glucose levels, even below the diagnostic level of GDM with adverse maternal and offspring outcomes. Based on the increasing risk of adverse maternal and offspring outcomes across 2-hour plasma glucose categories with a change of ∼1 mmol/L, we consider a difference of 1 mmol/L in 2-hour plasma glucose after OGTT between the intervention and control group as clinically relevant. We also used the observed standard deviation (1 SD = 1.3 mmol/L) in 2-hour plasma glucose after OGTT in the HAPO study for the sample size calculations. Calculation of the sample size for a two-sided t-test to detect a difference of 1 mmol/L between the groups, using an SD of 1.3 mmol/L, a power of 0.90, and a significance level of 0.05, yields 37 participants in each group in gestational week 28. To allow for an expected exclusion from the study due to not conceiving within the study period (∼50%)(64) yielding 74 per group, further drop-out during the study period (10-20%), yielding 93 per group, and to increase statistical power for secondary analyses, we initially wanted to include 260 participants in the trial.

However, we terminated the inclusion of new participants at 167 participants since we had reached 47 participants in each group who were pregnant in gestational week 12. With this number of participants, we foresee that we will have at least 37 participants in each group in gestational week 28, allowing for up to 20% dropout during pregnancy. We expect more participants who are already included to become pregnant in the upcoming period, which will increase the number of pregnant participants.

### Statistical analyses

The primary analysis will be done according to the ‘intention to treat’ principle, using all obtained data irrespective of participant adherence to the intervention and completeness of outcome measures. We plan to use linear mixed models (LMMs) to compare primary and secondary continuous outcome measures between groups, with time and group x time interactions as fixed effects variables, and subject as random factor.(65) Since no systematic baseline differences between the groups are expected in RCTs, means at baseline will be constrained to be equal in the LMMs. We will report estimates with corresponding 95% confidence intervals and p-values for differences between the intervention group and the control group. We will check the normality of residuals by visual inspection of QQ-plots and bootstrapping, transformations or non-parametric methods will be used in cases of non-normal model residuals. For the primary outcome measure, we will consider a p-value < 0.05 as statistically significant. For the secondary outcome measures, p-values < 0.01 will be considered statistically significant, due to multiple comparisons, and these analyses will be explorative. We will also perform per-protocol analyses: Participants with an average of ≥ 75 PAI per rolling week and adherence to TRE (as per definition above) during the preconception period will be included in the per-protocol analyses for all outcome measures. We will report additional results from all participants who were included in the trial, from the preconception period, irrespective of whether they became pregnant or not during the study period.

### Blinding

The study is not blinded as it is difficult to blind participants and treatment providers to behavioural intervention. However, baseline assessments are undertaken before randomisation.

### Monitoring

We do not expect any adverse effects in this study. If pregnant women are worried about foetal safety during exercise, we have experienced personnel available in the research group to monitor foetal heart rate during exercise sessions. The investigators are responsible for the documentation of any adverse or serious adverse events in the Case Report Form and the Serious Adverse Events Report Form, respectively. Participants are advised to contact the investigators if they have any unusual symptoms. All serious adverse events will be reported to the sponsor (NTNU) within 24 hours after the investigators have been informed of the event.

### Patient and public involvement

We have involved users in the planning of the study and will continue involving them in the implementation and dissemination. In the planning phase, we arranged a 1-hour digital workshop with users (reproductive-aged females with overweight/obesity), where we encouraged the audience to ask questions and give us feedback about relevant topics or issues related to participation. Regarding the challenges of long-term adherence, we use these feedbacks to find ways to incorporate exercise training and TRE into daily life.

### Ethics and dissemination

The Regional Committees for Medical and Health Research Ethics in Norway approved the study (REK, reference number 143756). The comparative analysis of the neonatal echocardiography data from this study with a corresponding group of neonates from mothers with normal BMI and no increased risk of GDM is also approved (REK reference number 67584). The work is conducted according to the Declaration of Helsinki and the ICMJE Recommendations for authorship. The participants sign an informed written consent before participating in the study and can at any time withdraw from the study without further explanation. Study specific ID numbers are used as participants’ identification. We ensure data quality by double data entry into an electronic CRF and treat the collected data following the General Data Protection Regulation. All protocol modifications are reported to REK. Upon completion of the study and finalization of the study report, we will submit the results for publication and/or in a publicly accessible database of clinical study results after anonymizing the data.

## Discussion

Based on a thorough literature search, the BTB study will be the first RCT to investigate the combined effects of TRE and exercise training, initiated before and continued throughout pregnancy, on cardiometabolic parameters in people at risk of GDM and their infants. We hypothesise that the combination of these two lifestyle interventions will induce an additive and clinically relevant improvement in maternal glucose tolerance, and potentially also in our secondary outcome measures in mothers and infants. As such, the initiation of lifestyle modification before pregnancy will provide a better platform for improved adherence and health outcomes, potentially breaking the intergenerational cycle of cardiometabolic disorders, and thereby reducing the risk of diabetes for future generations.

So far, there is limited data on the combination of TRE and exercise training in humans. Haganes and colleagues reported that the combination of TRE and HIIT in females with a BMI of ≥ 27 kg/m^2^ for 7 weeks significantly reduced HbA1c compared with a no-intervention control group and lead to greater losses in body weight, fat mass, and visceral fat area compared with either intervention alone.(26) Since the duration of the intervention is much longer in the BTB trial, there may be lower adherence to one or both intervention strategies. The possible reasons for lower adherence are that the participants may lack motivation for such a long time, find the intervention program boring and/or difficult, or develop physical symptoms that may hinder the participant to adhere to the intervention (especially during pregnancy). Combining motivational human interaction with digital interventions can increase engagement and the effectiveness of behaviour change interventions.(66) To improve adherence throughout the study period, we offer an individualised exercise regimen and provide encouragement, support, and monitor the participants regularly, both in person and over the phone.

The incidence and risk of obesity, insulin resistance, and GDM persists through generations.(67) To disrupt this intergenerational cycle, it is urgently necessary to develop and implement effective and practical lifestyle intervention strategies which can improve the cardiometabolic health outcomes of both mother and offspring. If the preconception lifestyle interventions implemented in this study lead to favourable outcomes and prove to be feasible and effective, it can pave the way for novel interventions that can be adopted in clinical practice during the preconception period, especially among those who are at risk of developing GDM.

## Author contributions

MAJS drafted the manuscript. TM, SAN, KÅS, ACI, TF, and SLF conceived and contributed to the design of the study and the plan for analyses. GR, MAJS, and HSS coordinate the study, perform measurements on test days, monitor participants, and supervise the exercise training. SAN performs the echocardiogram on the newborns. All authors provided feedback and approved the final manuscript.

## Data Availability

All data produced will be available online in the clinical trial registry.

https://classic.clinicaltrials.gov/ct2/show/NCT04585581?term=NCT04585581&recrs=d&cntry=NO&city=Trondheim&draw=2&rank=1

## Acknowledgments

The authors wish to thank all the participants for their contribution. We also thank the other members of the research team, Elisabeth Axe and Hilde Lund, who contributed to the execution of the BEFORE THE BEGINNING Study. The equipment and lab facilities for cardiorespiratory fitness testing is provided by NeXt Move, Norwegian University of Science and Technology (NTNU), and the clinical measurements are obtained at the Clinical Research Facility, St. Olavs Hospital. We would also like to thank the midwives at the Women and Children’s Centre, St. Olavs Hospital for the collection of samples related to birth. eFORSK, a stand-alone form-based information and communications technology solution for electronic collection of data, developed by Central Norway Regional Health Authority is used for sending invitations to the study.

