## Supplementary figures and images for "A randomised controlled trial of preconception lifestyle intervention on maternal and offspring health in people with increased risk of gestational diabetes: study protocol for the BEFORE THE BEGINNING trial"

### Figure 1

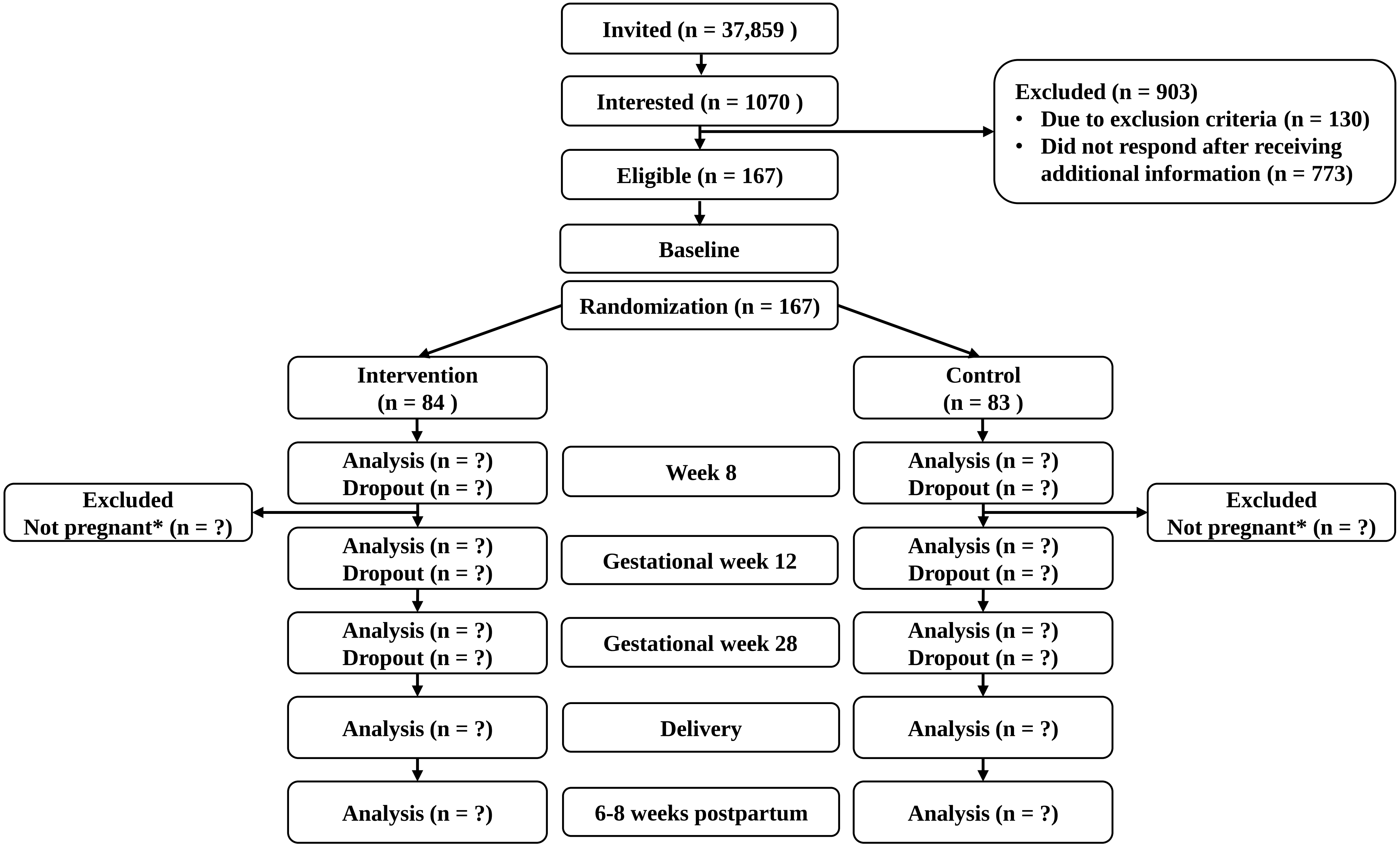

### Figure 2

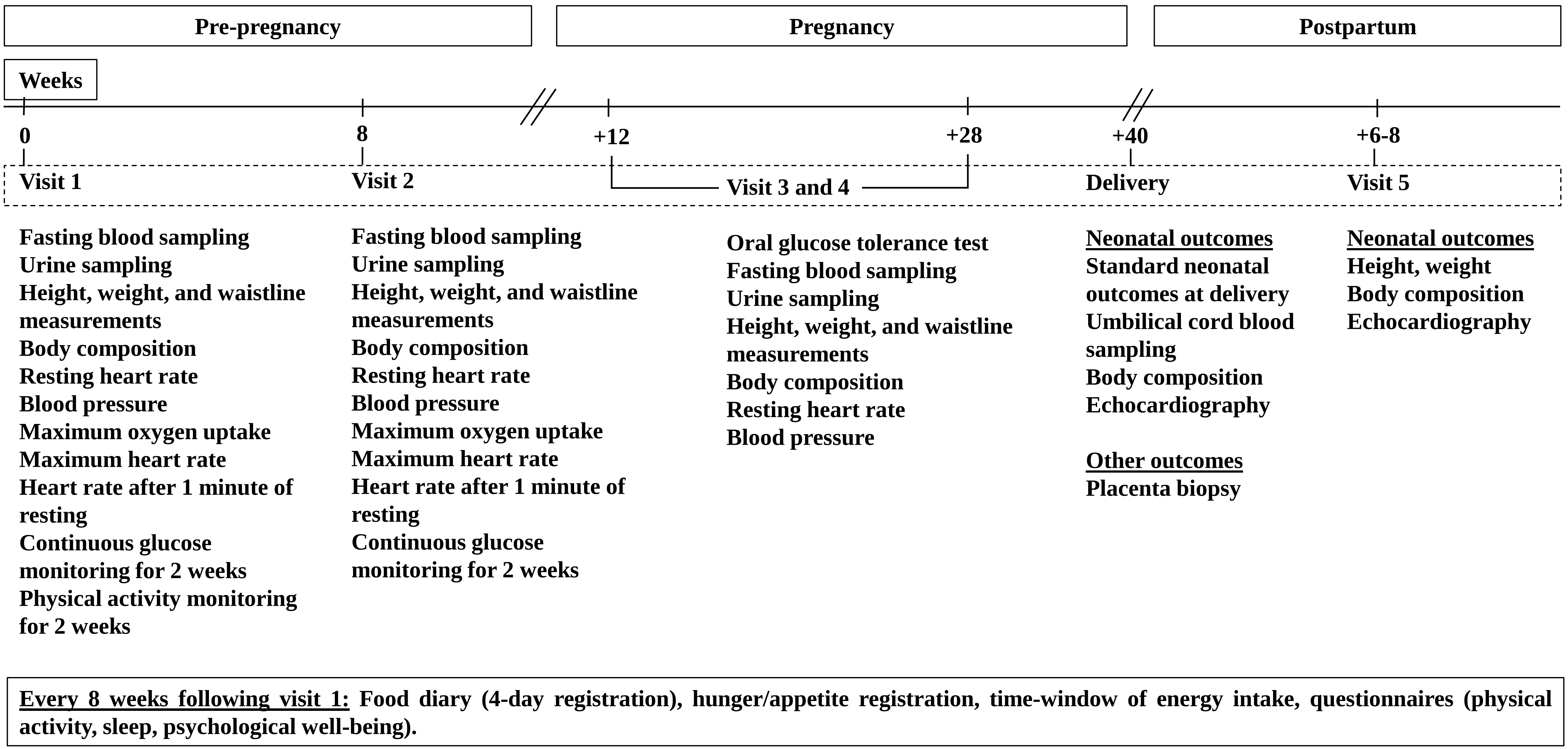
